# Barriers and Facilitators to Community Pharmacist-Provided Injectable Naltrexone for Formerly Incarcerated Individuals During Community Reentry in Wisconsin

**DOI:** 10.1101/2024.09.13.24313637

**Authors:** Jason S. Chladek, Michelle A. Chui

**Affiliations:** Grossman School of Medicine, New York University, 180 Madison Ave, New York, NY, 10016; Division of Social and Administrative Sciences, University of Wisconsin-Madison School of Pharmacy, 777 Highland Ave, Madison, WI, 53705; Sonderegger Research Center, University of Wisconsin-Madison School of Pharmacy, 777 Highland Ave, Madison, WI, 53705

**Keywords:** Community pharmacists, MOUD, injectable naltrexone, formerly incarcerated, OUD, reentry

## Abstract

Medications for opioid use disorder (MOUD), including injectable naltrexone, are a key component in the treatment of opioid use disorder (OUD). These medications are especially important for individuals transitioning out of correctional facilities and back into their communities, as individuals receiving MOUD are 85% less likely to die due to drug overdose in the first month post-release and have a 32% lower risk of rearrest. Unfortunately, few formerly incarcerated individuals have access to MOUD upon reentry, incurring a 40-fold greater likelihood of overdose following release compared to the general population. While 84% of Wisconsin jails offering MOUD offer naltrexone, less than half provide linkage to community treatment for reentering individuals. In Wisconsin, community pharmacists have the authority to provide naltrexone injections. However, they have not been explored as a resource for improving access to this medication for formerly incarcerated individuals. As a first step, the goal of this study was to understand the barriers and facilitators impacting access to community pharmacist-provided injectable naltrexone for this patient population during community reentry period. The researcher conducted semi-structured interviews with 18 individuals representing five stakeholder groups. Deductive and inductive content analysis were used to identify barrier and facilitator categories across the five levels of the Socioecological Model. Overall, participants discussed factors at every level, and many barriers and facilitators confirmed findings from existing literature focused on MOUD access for formerly incarcerated individuals. Participants also identified factors more specific to community pharmacies, including 1) lack of interagency collaboration between pharmacists, prescribers, and correctional facilities and 2) lack of awareness of community pharmacist-provided MOUD services. Future research should explore interventions to address the barriers identified in this study and improve connections between community pharmacists and formerly incarcerated individuals. This work can help ensure that these individuals are given the chance to successfully reintegrate into society.

## Introduction

The opioid epidemic is a major public health issue in the United States. More than three million citizens suffer from opioid use disorder (OUD), a problematic pattern of opioid use leading to health problems and/or social distress.^1–3^ Specifically, Wisconsin has been significantly impacted by this problem, with opioid overdose deaths increasing 900% from 1999 to 2019. In 2022 alone, there were 1,464 opioid-related deaths in the state.^4–5^

OUD is highly prevalent among individuals involved in the criminal justice system. In 2020, the Wisconsin Department of Corrections (DOC) reported 325 deaths among those admitted to probation and 276 among those released from prison.^6^ Medications for opioid use disorder (MOUD), which include methadone, buprenorphine, and naltrexone, are a key component in the treatment of OUD, and are especially important for individuals transitioning out of correctional facilities and back into their communities.^7^ Formerly incarcerated individuals receiving MOUD are 85% less likely to die due to drug overdose in the first month after release and have a 32% lower risk of rearrest.^8^

Unfortunately, few formerly incarcerated individuals are able to access sustainable MOUD treatment upon community reentry, missing a critical tool for rehabilitation and incurring a 40-fold greater likelihood of opioid overdose following release compared to the general population.^9^ Previous work has shown that in individuals who are released with doses of MOUD, less than half continue use in the community.^10–12^ In Wisconsin, only 47.7% of jails provided those being released with a community link to MOUD.^13–14^ The Substance Abuse and Mental Health Services Administration (SAMHSA) reports that 40-50% of these individuals are rearrested within a year of release, and 75% relapse to opioid use within three months.^15^ Furthermore, lack of access to MOUD during reentry is tied to racial and ethnic disparities, as Black, Hispanic, and Latine populations are disproportionately impacted.^16–17^ Overall, there is a clear need to improve access to MOUD for formerly incarcerated individuals during community reentry. The volume of research in progress shows that more professionals are recognizing this need, but this work remains limited.^18^

While current research efforts are limited, there are certain components of existing interventions and programs that show promise. For example, the success of mobile treatment demonstrates that an accessible location for MOUD treatment can facilitate access. Another unexplored resource that could provide an accessible location is community pharmacies.^19^ Community pharmacists are not only considered more accessible than other healthcare providers, but 96.5% of the U.S. population lives within 10 miles of a community pharmacy.^20–21^

Wisconsin community pharmacists have had the authority to administer long-acting injectable naltrexone treatments since 2019.^22^ For formerly incarcerated individuals, injectable naltrexone is associated with improved treatment retention, reduced healthcare utilization, reduced rates of reincarceration, reduced opioid relapse, and improved medication adherence. Additionally, injectable naltrexone is long-lasting and has a decreased risk of abuse potential, making it widely accepted and used among justice-impacted individuals.^23^

Long-term, improving connections between formerly incarcerated individuals and community pharmacists can help increase access to MOUD during the community reentry period. As a first step, the goal of this study is to understand the exiting barriers and facilitators to this care. While previous work has examined barriers and facilitators to MOUD for formerly incarcerated individuals, as well as barriers and facilitators faced by community pharmacists in providing these services, community pharmacist-provided injectable naltrexone has not been explored for this particular population.^22,24–35^ This study will utilize semi-structured interviews with various stakeholder groups to comprehensively identify existing barriers and facilitators impacting the availability, access, and use to these services for formerly incarcerated individuals.

## Materials and Methods

### Participants and sampling

Participants were recruited for individual semi-structured interviews between September 2023 and January 2024. Study participants were recruited if they were identified as potential stakeholders in transitions of care for formerly incarcerated individuals with opioid use disorder during the community reentry process. Individuals fell within one of five different stakeholder groups: 1) MOUD prescribers with experience providing care for formerly incarcerated patients, 2) community pharmacists with experience administering naltrexone injections for formerly incarcerated patients, 3) professionals working in a correctional setting with experience assisting formerly incarcerated individuals with OUD during reentry planning, 4) professionals working for a community organization or non-profit with experience assisting formerly incarcerated individuals with OUD during reentry planning, and 5) individual patients with a history of incarceration and using injectable naltrexone for OUD treatment OR a family member/caregiver of an individual with a history of incarceration and using injectable naltrexone for OUD treatment.

Participants from all five stakeholder groups were 18 years of age or older, able to speak and understand English, and residing in Wisconsin. The goal of recruiting individuals from different stakeholder groups was to comprehensively understand the barriers and facilitators to accessing community pharmacist-provided injectable naltrexone from multiple perspectives. This approach was also used to help ensure that barriers and facilitators from every level of the Socioecological Model were discussed. Individual patients and family members/caregivers were combined into one category, as it was anticipated that both groups would offer similar perspectives. Additionally, patients and family members/caregivers were not recruited from the same family.

The lead researcher had established relationships with several primary health clinics, pharmacies, and community organizations throughout Wisconsin, which were leveraged to identify and recruit participants. Initial recruitment was limited, especially concerning correctional staff and formerly incarcerated patients, so snowball sampling was utilized to identify additional participants who fit the inclusion criteria. In total, 18 participants were recruited, as shown in Table 1. This study was deemed exempt by the (XXXX) Institutional Review Board.

**Table 1.** Participants by stakeholder group.

| Role | No. of participants |
| --- | --- |
| MOUD prescribers | 4 |
| Community pharmacists | 3 |
| Correctional staff | 4 |
| Community organization or non-profit staff | 4 |
| Individual patients OR family members/caregivers | 3 |

### Procedures

All potential participants were informed of the study and invited to participate via email. After indicating an interest in participating, they were emailed an informational sheet about the project and interviews were scheduled. The informational sheet was reviewed by the researcher on the call prior to the start of the interview, after which verbal consent to participate was obtained. The lead researcher emphasized that there was no obligation to participate, and participation was voluntary and could be stopped at any time. All interviews were conducted via Zoom by the lead researcher. Interviews were audio recorded to help facilitate transcription and took 45 minutes to 1 hour. After the interview, participants were sent a five-minute demographic survey, which was returned to the researcher via email. Participants were compensated with $60 gift cards after completion of the interview and survey.

The lead researcher conducted semi-structured interviews to identify the barriers and facilitators to community pharmacist-provided injectable naltrexone for formerly incarcerated individuals during community reentry. Two interview guides were created by the researcher that aligned with 1) providers, pharmacists, or staff and 2) patients, family members, or caregivers. The researcher anticipated that the use of community pharmacist-provided injectable naltrexone by formerly incarcerated individuals during reentry was limited, and not every participant would have direct experience with coordinating, providing, or receiving these services. As a result, the interview guides included questions for those with or without direct experience. Participants were first asked whether or not they had experience coordinating, providing, or receiving community pharmacist-provided naltrexone injections. If not, participants were asked to discuss anticipated barriers and facilitators based on their perceptions and/or previous experiences with reentry planning and using community pharmacies for healthcare services. Additionally, the researcher did not ask about any experiences related to drug abuse or addiction outside of access to treatment, and participants were told that they did not have to answer any questions or share any details they were uncomfortable discussing.

### Data coding and analysis

The interviews were transcribed verbatim, de-identified, and verified for accuracy. All participants were assigned an ID number based on their stakeholder group. Transcripts were entered into NVivo, a qualitative data software package (released in March 2020).^36^ The researchers performed deductive and inductive qualitative content analysis as outlined Elo & Kyngäs.^37^ Both deductive and inductive approaches were used, as there is some previous knowledge on the barriers and facilitators that impact MOUD access for formerly incarcerated individuals, as well as factors impacting community pharmacists’ abilities to implement injectable naltrexone services. However, knowledge related specifically to community pharmacist-provided injectable naltrexone for formerly incarcerated individuals is highly limited.

First, the lead researcher developed a categorization matrix based on the five domains of the Socioecological Model, visualized in Table 2. The Socioecological Model, as shown in Figure 1, is a multilevel model that conceptualizes factors impacting health behaviors and outcomes, as well as the interactions between these factors. It also supports the idea that behaviors both affect and are affected by various contexts.^38–41^ The Model has been used extensively in public and population health efforts, including identifying barriers and facilitators to healthcare services. It has also been applied to studies focused on vulnerable populations, including individuals with a history of incarceration and/or substance use disorders.^32,40,42–50^

**Table 2.** Categorization matrix for content analysis.

|  | Public Policy | Community | Organizational | Interpersonal | Individual |
| --- | --- | --- | --- | --- | --- |
| Barriers |  |  |  |  |  |
| Facilitators |  |  |  |  |  |

**Figure 1.**
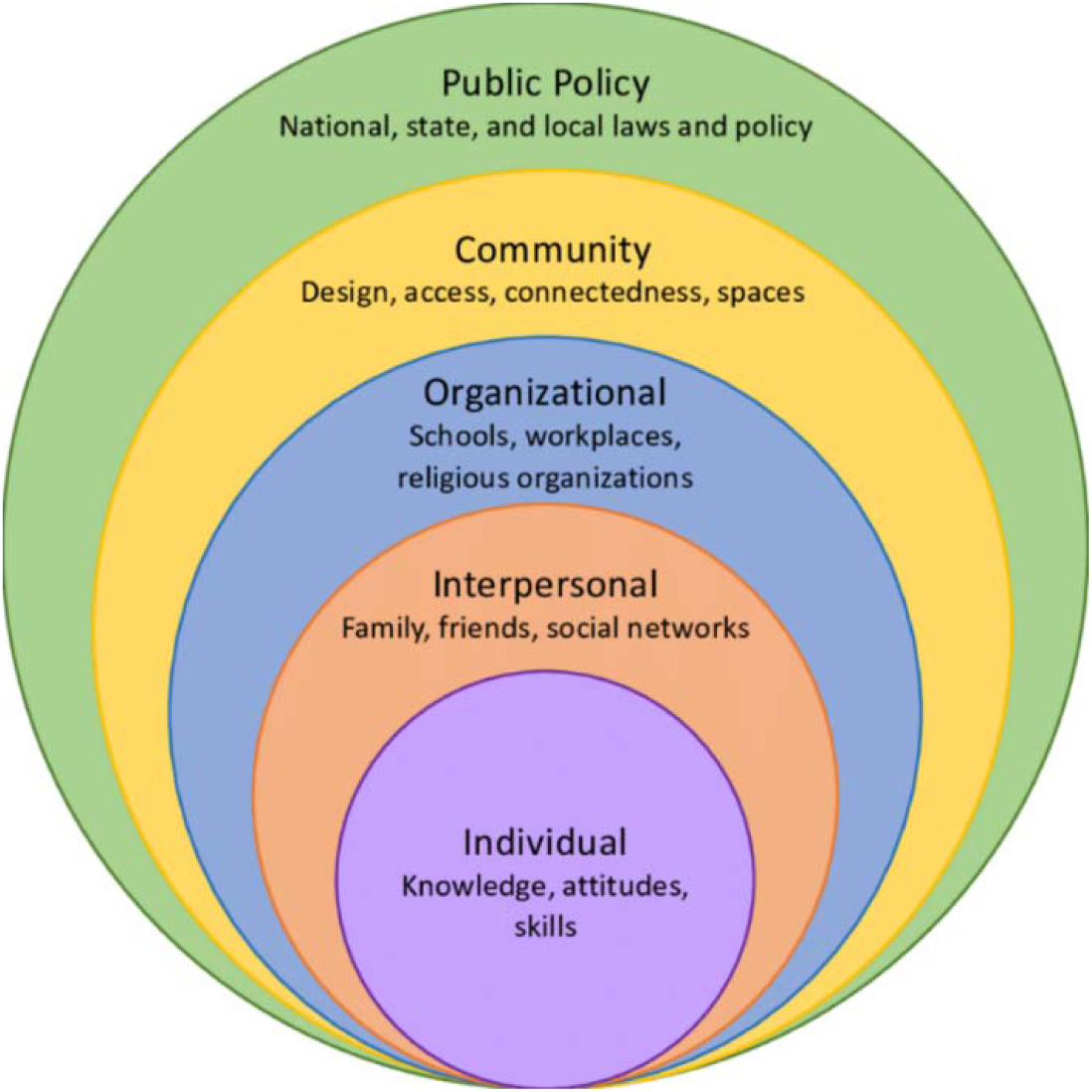
The Socioecological Model^41^.

The researchers applied a deductive approach by analyzing the transcripts line-by-line and coding the data according to the matrix. Factors were categorized as a barrier or facilitator depending on whether the participant was talking about availability, access, and/or use of community pharmacist-provided injectable naltrexone being hindered or supported by that specific factor. To determine the level of the Socioecological Model, the researchers evaluated the context of each factor. For example, if a participant stated that they did not have personal access to a car or mode of transportation, this would have been coded to the individual level. However, if a participant stated that their neighborhood did not have reliable public transportation, this would have been coded to the community level. Any discrepancies were resolved during discussions between both coders.

Next, an inductive approach was used to group the data within each domain and create higher order categories. Development of categories was supported and confirmed through discussions between researchers. Any ambiguities were also addressed during these discussions. The lead researcher created a comparative analysis table to highlight which stakeholder groups discussed each of the categories (for both barriers and facilitators). This was used to compare responses between groups and highlight convergence and divergence across the different stakeholders. Finally, representative quotes were selected for each of the categories. Overall, this process was guided by the four-dimension criteria of qualitative research.^51^

## Results

In total, 18 stakeholders participated in an interview. Participant demographics are outlined in Table 3. The Socioecological Model offered a framework for conceptualizing the factors impacting access to community pharmacist-provided naltrexone injections for formerly incarcerated individuals during the community reentry period.^38–41^

**Table 3.** Participant demographics.

|  | **MOUD**<br>**prescribers**<br>**(n=4)** | **Community**<br>**pharmacists**<br>**(n=3)** | **Correctional**<br>**staff**<br>**(n=4)** | **Community**<br>**organization**<br>**staff**<br>**(n=4)** | **Patients,**<br>**family,**<br>**caregivers**<br>**(n=3)** | **Total**<br>**(n=18)** |
| --- | --- | --- | --- | --- | --- | --- |
| **Age** | 40.25 | 37.33 | 31.00 | 39.50 | 44.67 | 38.28 |
| **Gender** |  |  |  |  |  |  |
| Male | 0 (0%) | 3 (100%) | 0 (0%) | 1 (25%) | 1 (33.33%) | 5 (27.78%) |
| Female | 4 (100%) | 0 (0%) | 4 (100%) | 3 (75%) | 2 (66.67%) | 13 (72.22%) |
| Other | 0 (0%) | 0 (0%) | 0 (0%) | 0 (0%) | 0 (0%) | 0 (0%) |
| **Ethnicity** |  |  |  |  |  |  |
| Hispanic/Latino | 1 (25%) | 0 (0%) | 0 (0%) | 1 (25%) | 0 (0%) | 2 (11.11%) |
| Not Hispanic or Latino | 3 (75%) | 3 (100%) | 4 (100%) | 3 (75%) | 3 (100%) | 16 (88.89%) |
| **Race** |  |  |  |  |  |  |
| White | 4 (100%) | 3 (100%) | 4 (100%) | 4 (100%) | 3 (100%) | 18 (100%) |
| Black/African American | 0 (0%) | 0 (0%) | 0 (0%) | 1 (25%) | 0 (0%) | 1 (5.56%) |
| Asian | 0 (0%) | 0 (0%) | 0 (0%) | 0 (0%) | 0 (0%) | 0 (0%) |
| Native Hawaiian/Pacific Islander | 0 (0%) | 0 (0%) | 0 (0%) | 0 (0%) | 0 (0%) | 0 (0%) |
| American Indian/Alaska Native | 0 (0%) | 0 (0%) | 0 (0%) | 0 (0%) | 0 (0%) | 0 (0%) |
| Other | 0 (0%) | 0 (0%) | 0 (0%) | 0 (0%) | 0 (0%) | 0 (0%) |
| **Educational Level** |  |  |  |  |  |  |
| Less than high school | 0 (0%) | 0 (0%) | 0 (0%) | 0 (0%) | 0 (0%) | 0 (0%) |
| High school or equivalent | 0 (0%) | 0 (0%) | 0 (0%) | 0 (0%) | 2 (66.67%) | 2 (11.11%) |
| Some college, no degree | 0 (0%) | 0 (0%) | 0 (0%) | 0 (0%) | 0 (0%) | 0 (0%) |
| Associate or Bachelor | 0 (0%) | 1 (33.33%) | 3 (75%) | 2 (50%) | 1 (33.33%) | 7 (38.89%) |
| Master or above | 4 (100%) | 2 (66.67%) | 1 (25%) | 2 (50%) | 0 (0%) | 9 (50.00%) |

For each level of the Socioecological Model, categories related to barriers and facilitators were distinguished, as displayed in Table 4. Table 5 and 6 show the comparative analysis for which stakeholder groups discussed each barrier or facilitator category, respectively. Overall, although participants belonged to different stakeholder groups and were speaking from different vantage points, many of their responses overlapped. As a result, the categories below pertain to the whole sample, and variations or nuances between stakeholder groups are discussed where applicable. Tables 7 and 8 highlight representative quotes for each of the categories, as well as the stakeholder group associated with the quote. All table colors correspond to the levels of the Socioecological Model as shown in Figure 1.

**Table 4.** Categories of barriers and facilitators to community pharmacist-provided naltrexone injections for formerly incarcerated individuals during community reentry.

| Barriers | Facilitators |
| --- | --- |
| ***Public Policy Level*** |  |
| • Cost of drug<br>• Cost of drug testing<br>• Prescription requirement | • OUD classification |
| ***Community Level*** |  |
| • Stigma<br>• Lack of interagency collaboration<br>• Lack of available prescribers/injectors | • Accessible pharmacy locations |
| ***Organizational Level*** |  |
| • Administrative constraints<br>• Lack of pharmacy advertising<br>• Inability of pharmacists to provide additional OUD services | • Flexibility of appointments<br>• Non-judgmental environment*<br>• Pharmacy hours* |
| ***Interpersonal Level*** |  |
| • Negative home/social environment | • Patient advocates/social support<br>• Patient-provider relationship<br>• Treatment reminders |
| ***Individual Level*** |  |
| • Lack of awareness<br>• Lack of insurance<br>• Lack of reliable transportation<br>• Lack of stable housing<br>• Competing priorities<br>• Medication side effects | • Having a plan and/or goals<br>• Readiness to change |

\*Categories labeled with an asterisk were discussed as both barriers and facilitators. However, they were placed under the domain they were most commonly identified as.

**Table 5.** Comparative analysis of barrier categories.

|  | **MOUD**<br>**prescribers**<br>**(n=4)** | **Community**<br>**pharmacists**<br>**(n=3)** | **Correctional**<br>**staff (n=4)** | **Organization**<br>**or non-profit**<br>**staff (n=4)** | **Patient,**<br>**family**<br>**member, or**<br>**caregiver**<br>**(n=3)** | **All**<br>**stakeholders**<br>**(n=18)** |
| --- | --- | --- | --- | --- | --- | --- |
| **Cost of drug** | 1 (25%) | 3 (75%) |  | 1 (25%) | 1 (33%) | 6 (33%) |
| **Cost of drug**<br>**testing** | 1 (25%) |  |  |  |  | 1 (6%) |
| **Prescription**<br>**requirement** | 1 (25%) | 1 (33%) |  | 2 (50%) |  | 4 (22%) |
| **Stigma** | 1 (25%) |  | 2 (50%) | 2 (50%) | 1 (33%) | 6 (33%) |
| **Lack of**<br>**interagency**<br>**collaboration** | 2 (50%) | 3 (75%) | 3 (75%) | 2 (50%) |  | 10 (56%) |
| **Lack of**<br>**available**<br>**prescribers or**<br>**injectors** | 3 (75%) |  | 1 (25%) | 1 (25%) |  | 5 (28%) |
| **Administrative**<br>**constraints** | 2 (50%) | 3 (75%) | 1 (25%) |  |  | 6 (33%) |
| **Lack of**<br>**pharmacy**<br>**advertising** | 1 (25%) | 2 (67%) |  |  |  | 3 (17%) |
| **Inability of**<br>**pharmacists to**<br>**provide**<br>**additional OUD**<br>**services** | 3 (75%) | 1 (33%) | 2 (50%) |  | 1 (33%) | 7 (39%) |
| **Negative home**<br>**or social**<br>**environment** | 1 (25%) |  |  |  | 1 (33%) | 2 (11%) |
| **Lack of**<br>**awareness** | 1 (25%) | 2 (67%) | 1 (25%) | 2 (50%) | 1 (33%) | 7 (39%) |
| **Lack of**<br>**insurance** | 3 (75%) | 2 (67%) |  | 4 (100%) | 2 (67%) | 11 (61%) |
| **Lack of reliable**<br>**transportation** | 3 (75%) | 3 (75%) | 4 (100%) | 3 (75%) | 2 (67%) | 15 (83%) |
| **Lack of stable**<br>**housing** | 1 (25%) | 1 (33%) | 1 (25%) | 1 (25%) |  | 4 (22%) |
| **Competing**<br>**priorities** | 1 (25%) |  | 1 (25%) | 1 (25%) |  | 3 (17%) |
| **Medication side**<br>**effects** |  | 1 (33%) | 1 (25%) |  | 1 (33%) | 3 (17%) |

**Table 6.**
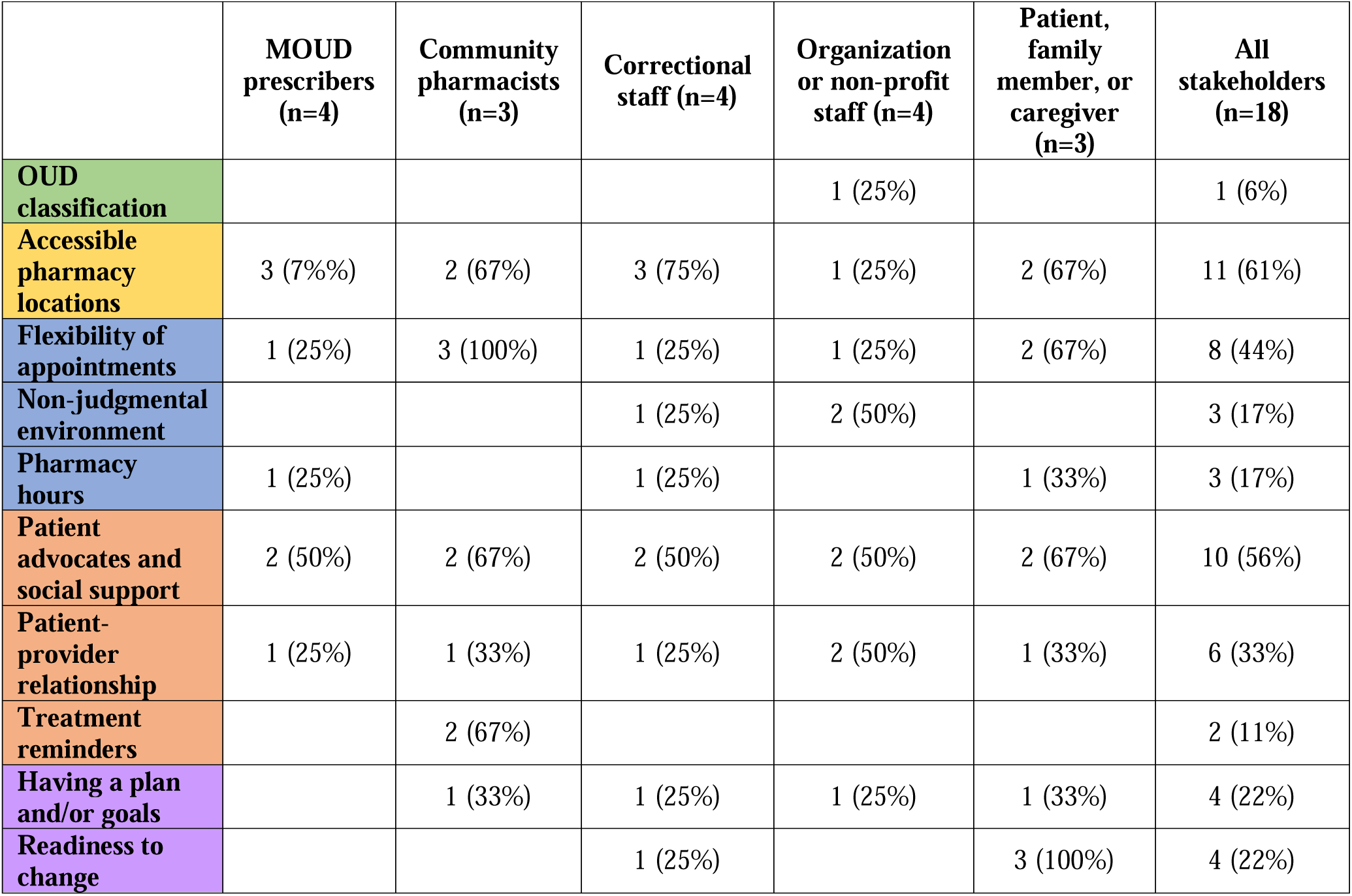
Comparative analysis of facilitator categories.

**Table 7.**
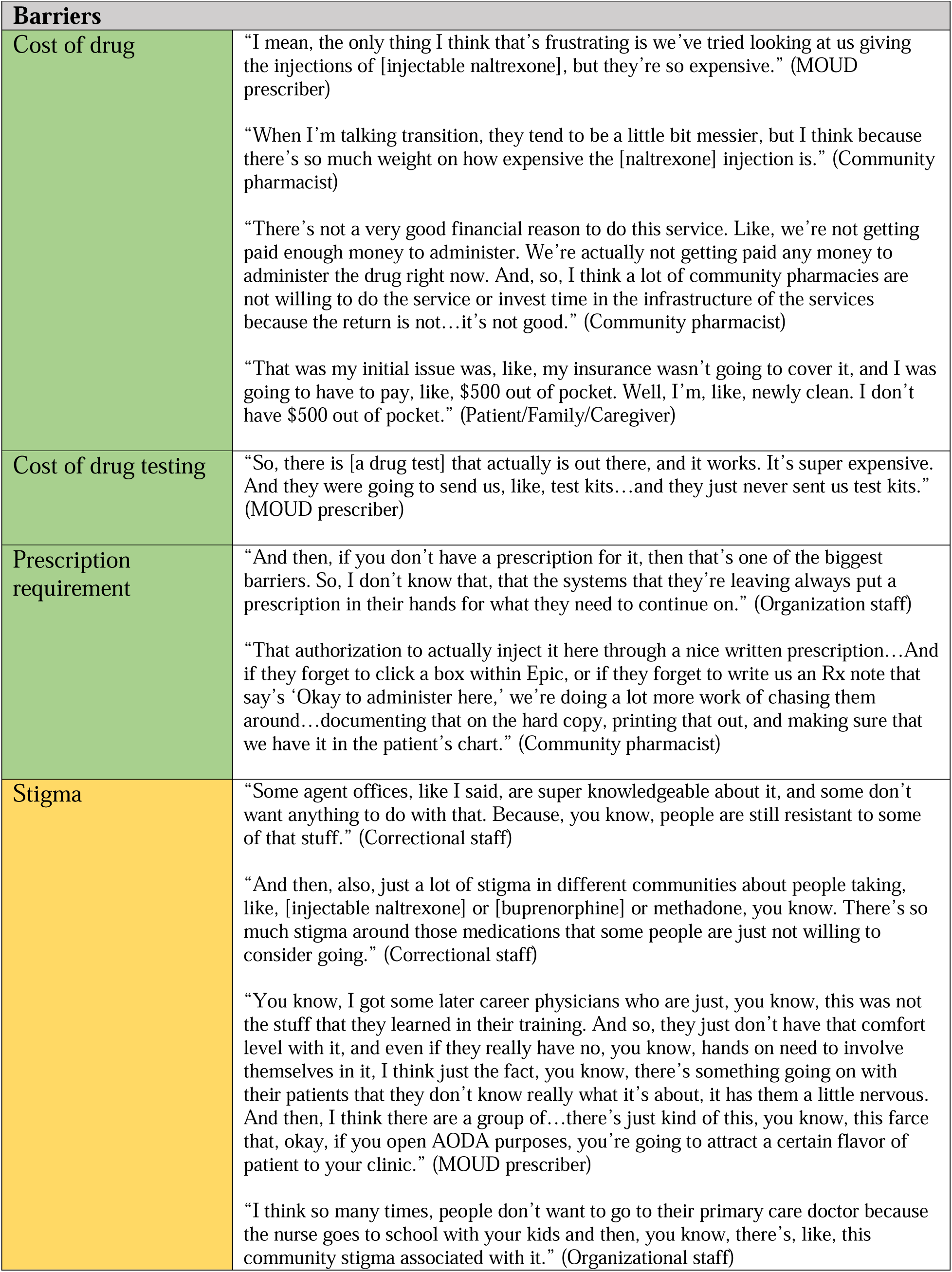
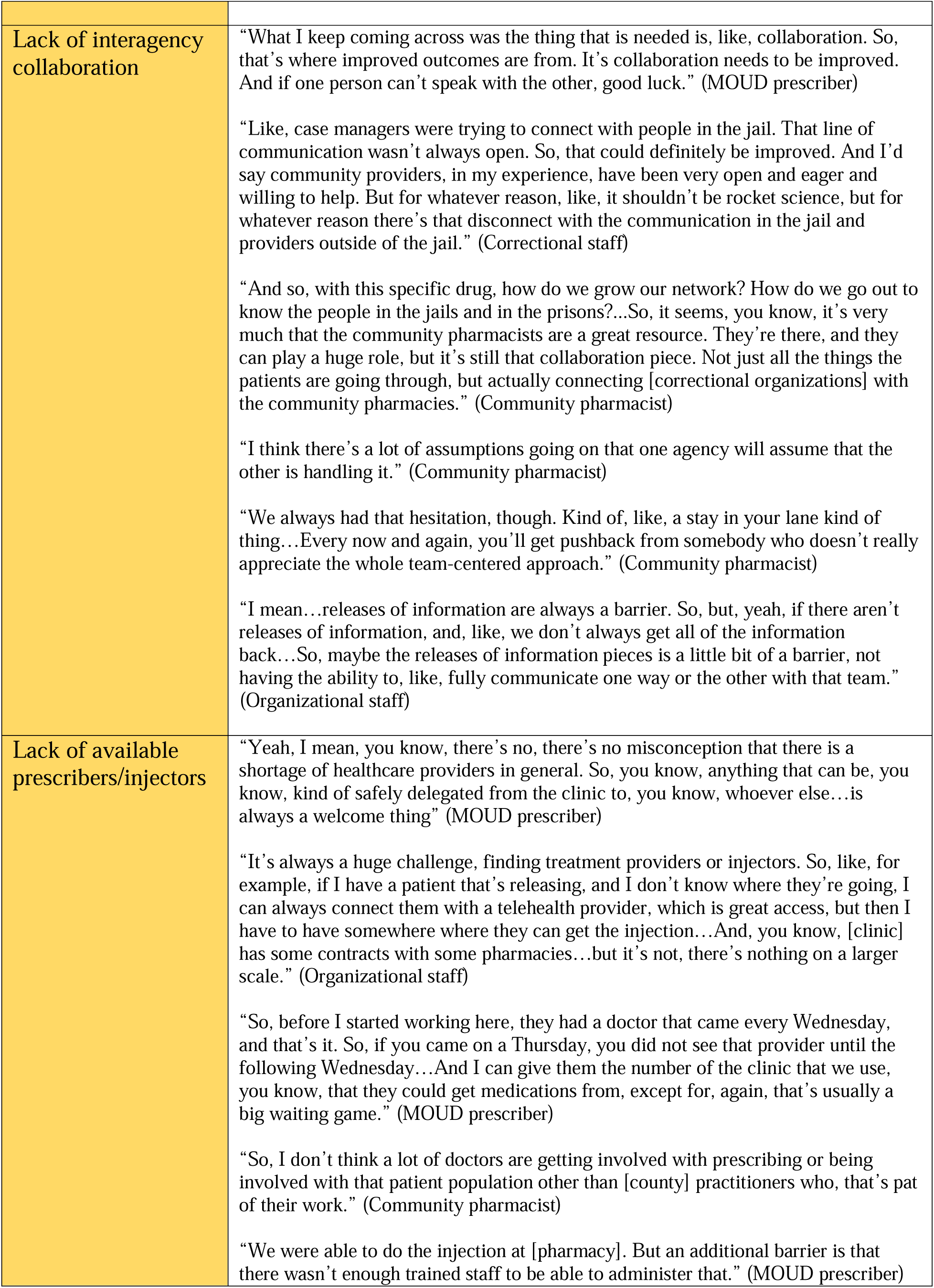
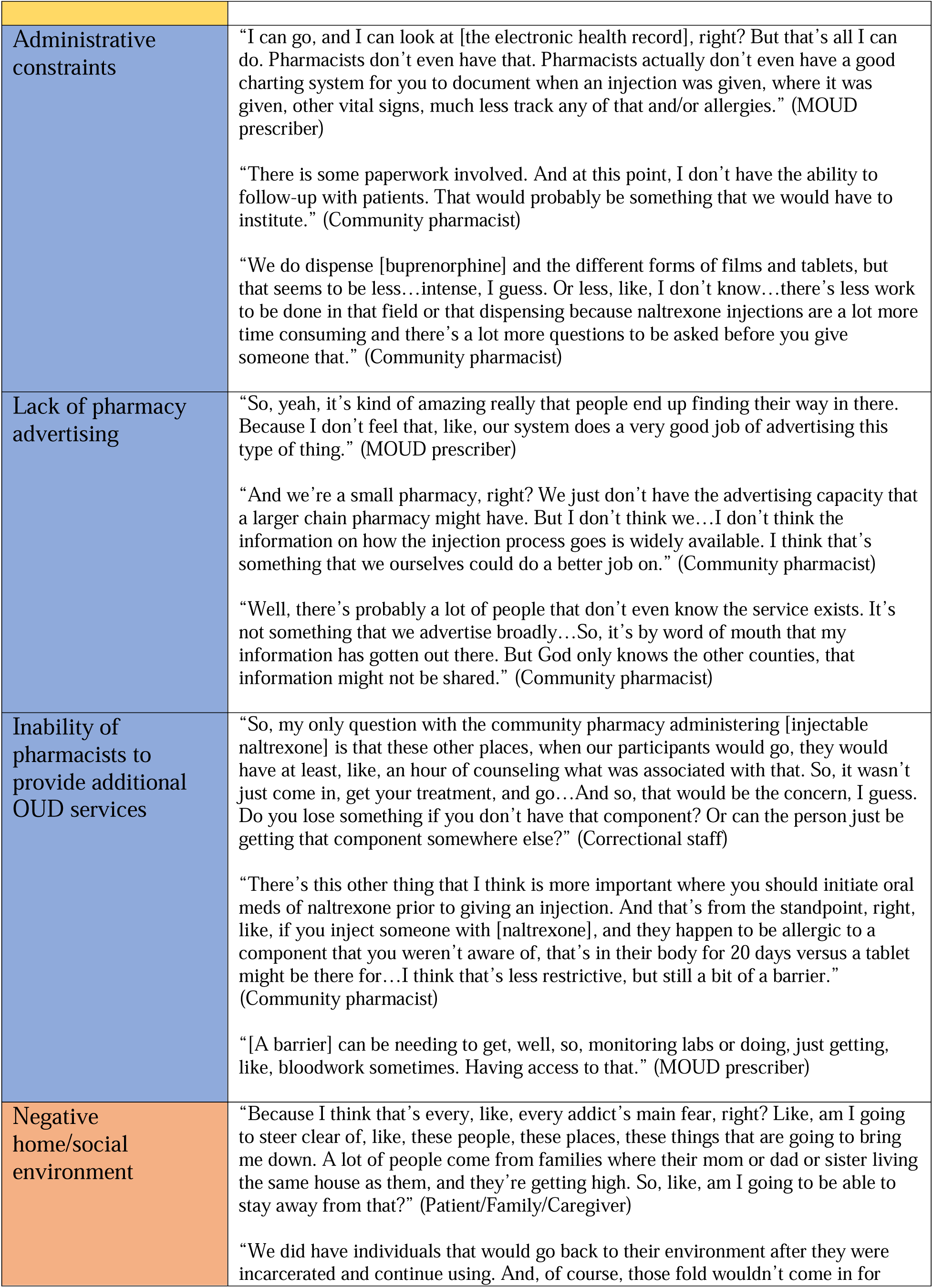
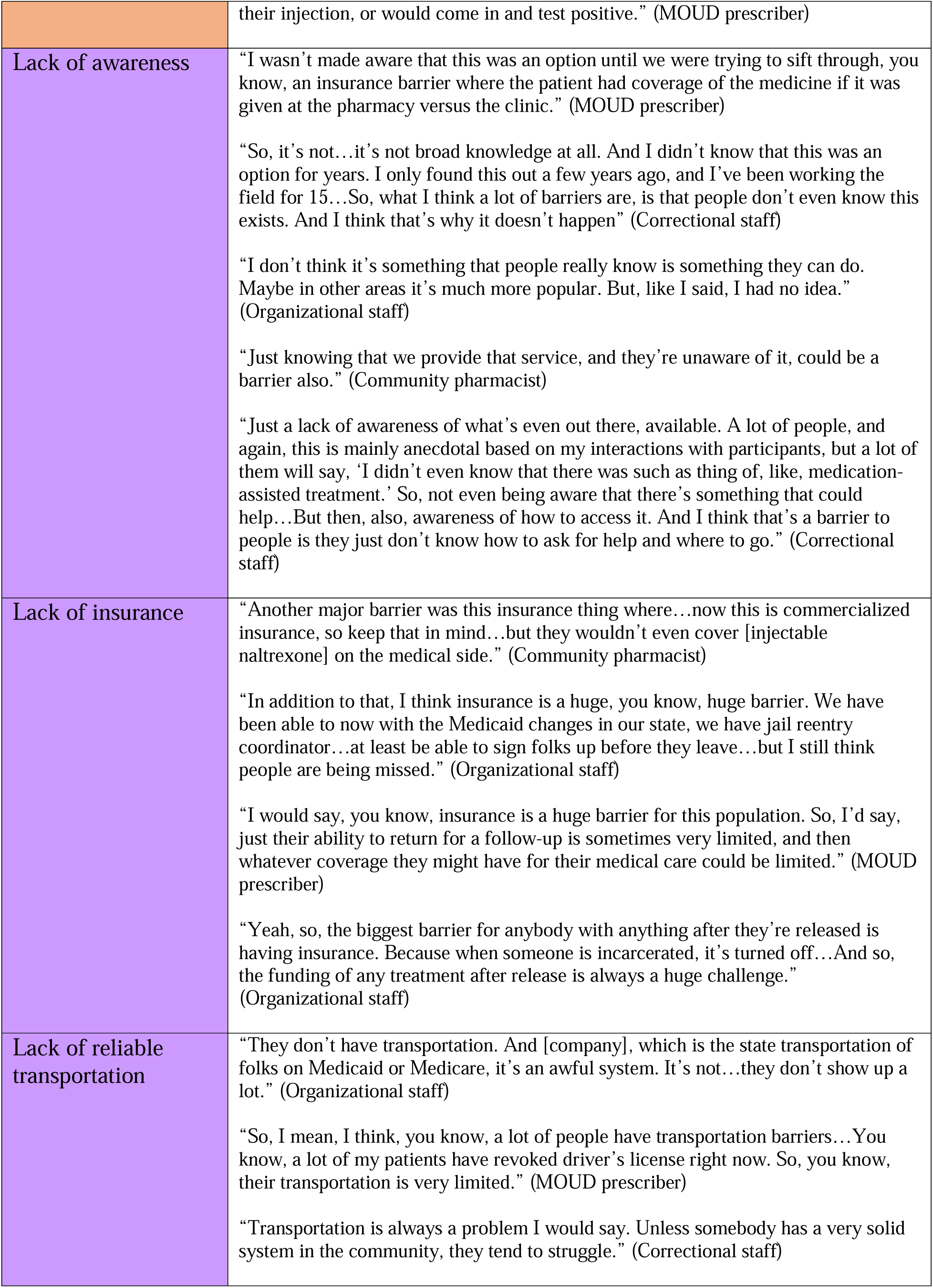
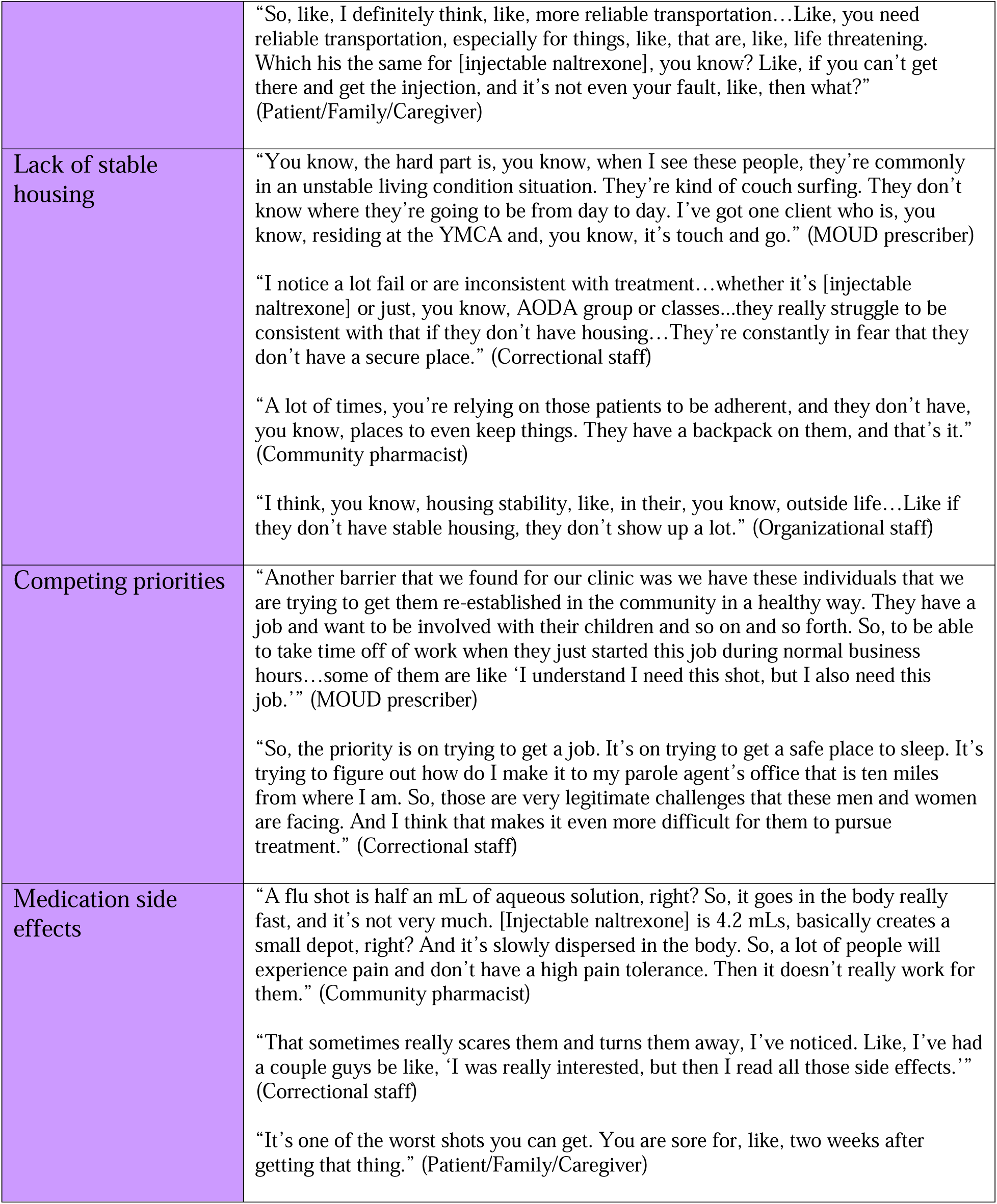
Representative quotes of barrier categories.

**Table 8.**
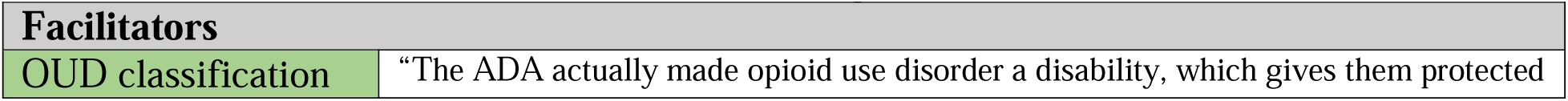
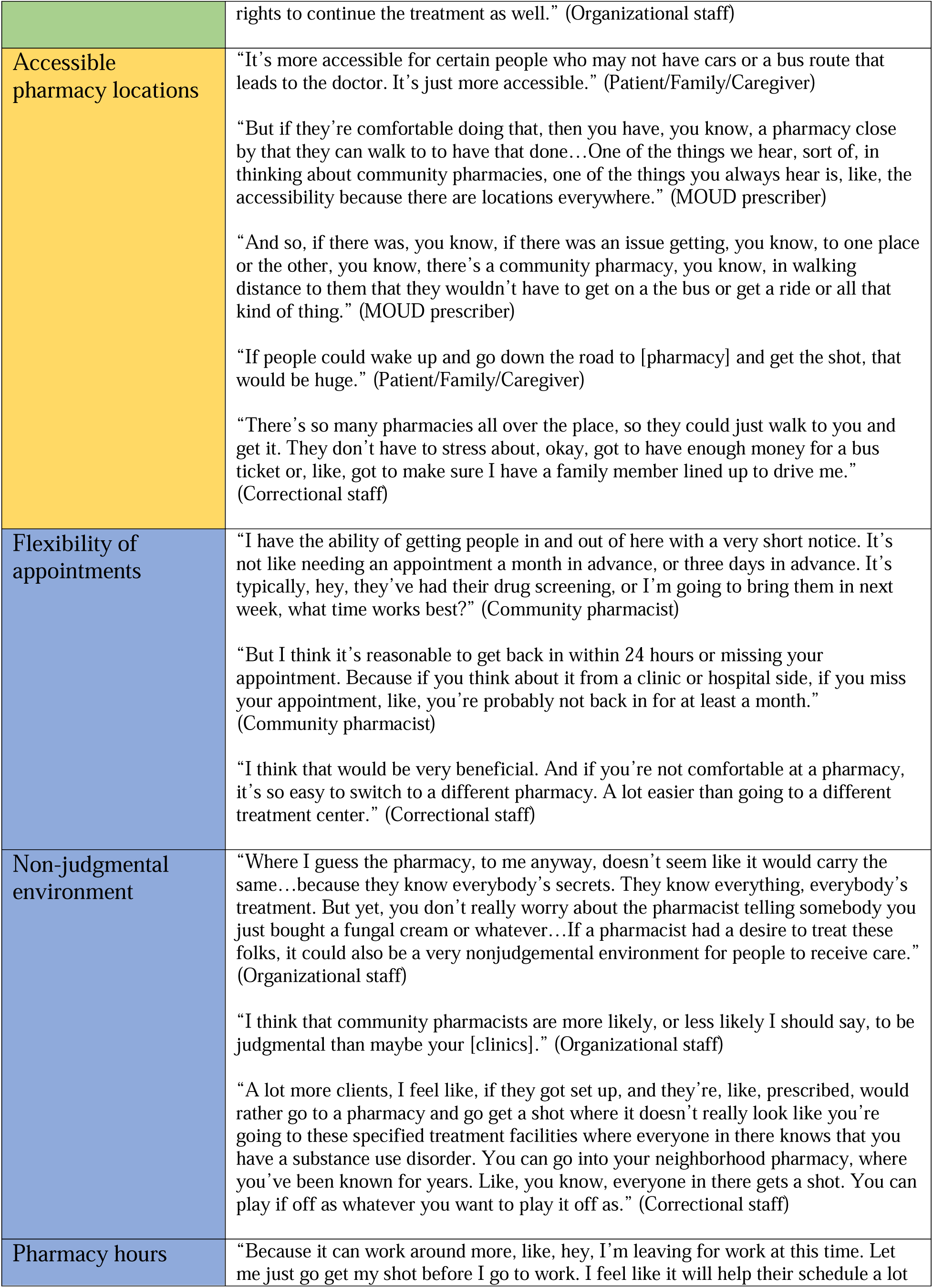
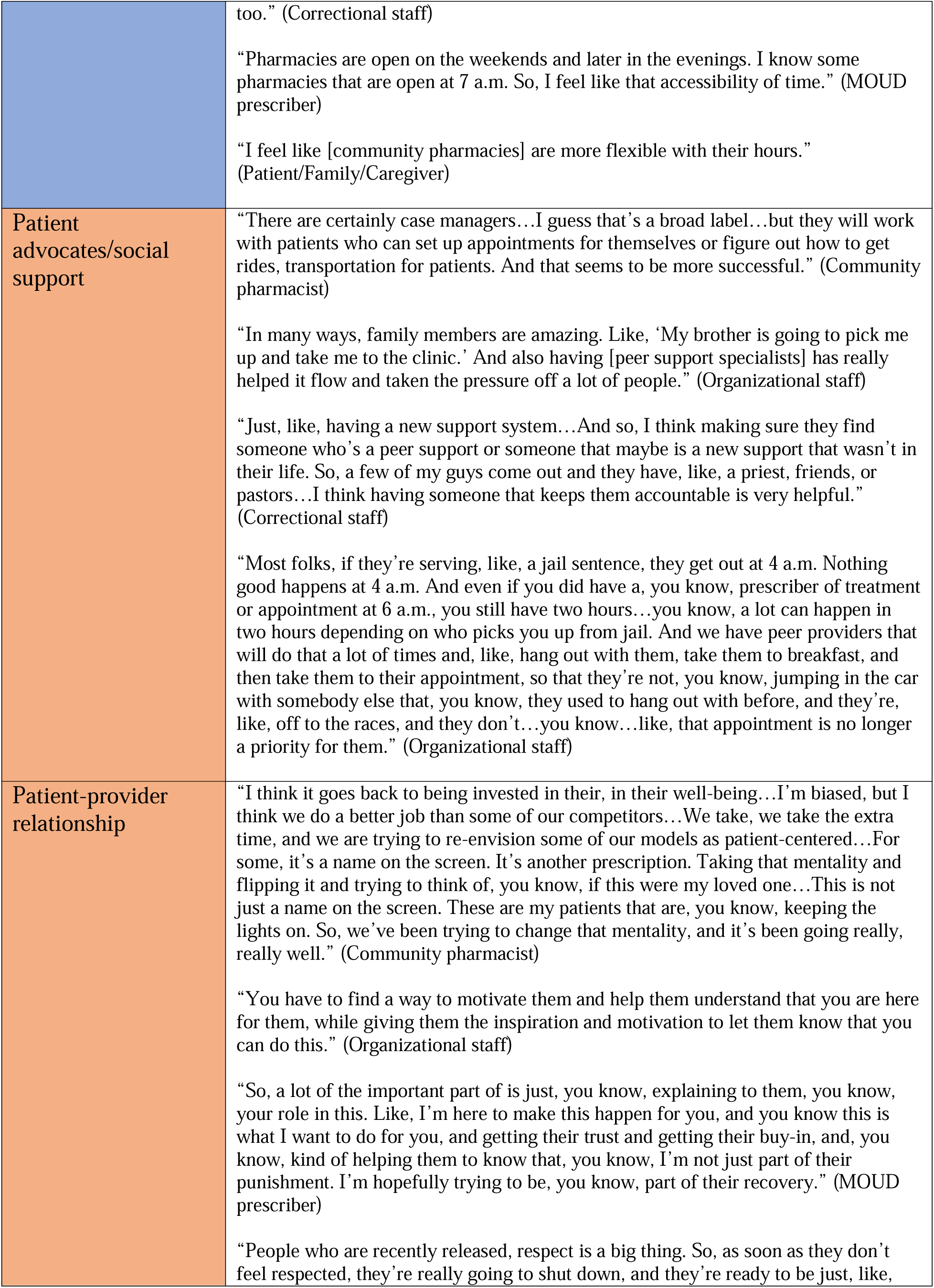
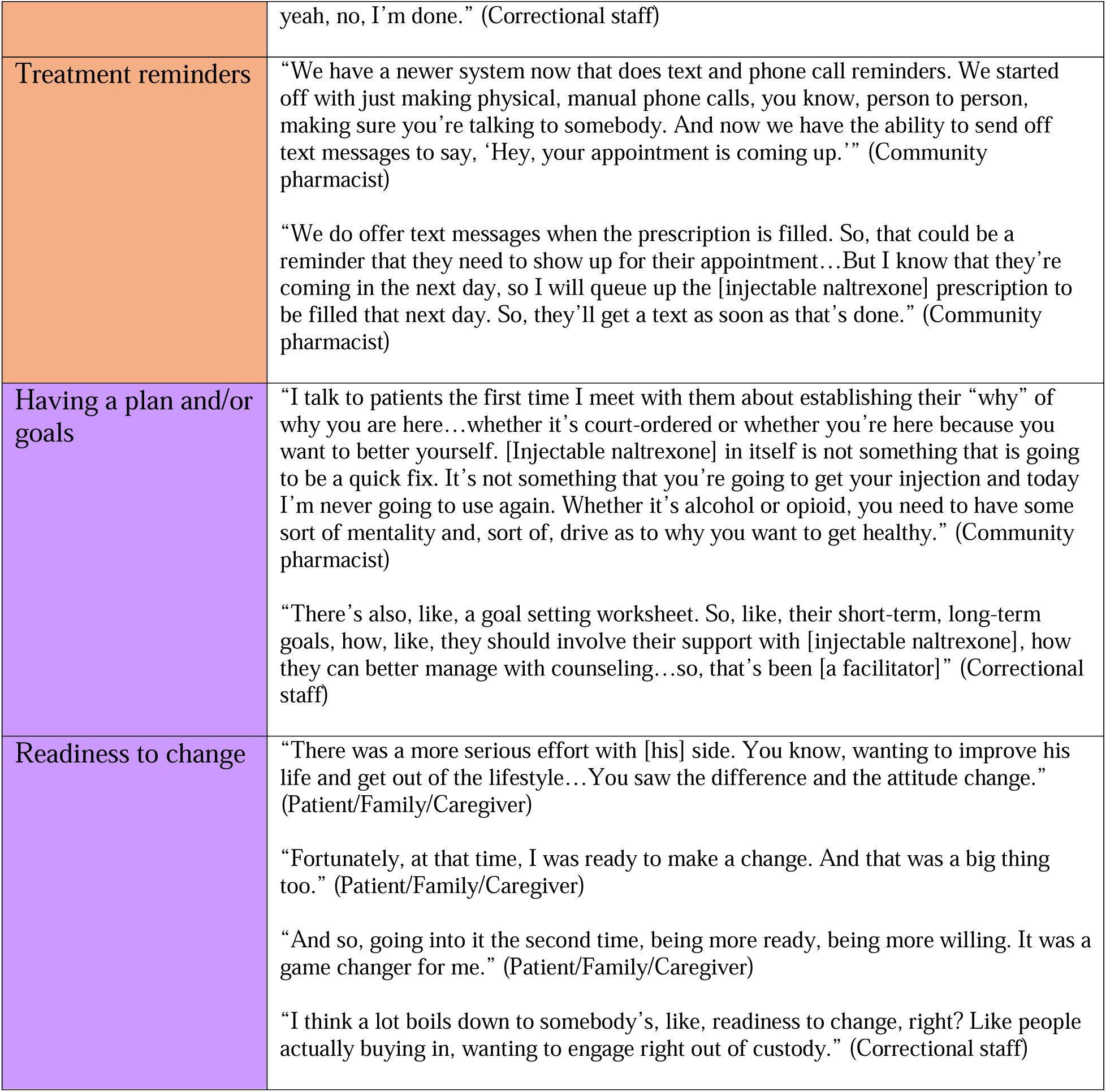
Representative quotes of facilitator categories.

### Public Policy Level

Overall, participants did not heavily discuss barriers and facilitators at the public policy level. Most of the discussion at this level centered around cost. For example, several participants said that the cost of naltrexone injections creates a major barrier. Few added that the cost of the medication greatly exceeds the reimbursement to the community pharmacies, so there is no financial incentive for them to provide the services or invest time in the infrastructure needed to provide injections. Overall, the expense can deter community pharmacists from providing the injections, limiting access to formerly incarcerated patients. Additionally, an MOUD prescriber added that the high cost of drug testing can create similar barriers, especially if community pharmacists take on the responsibility of testing patients prior to injections. Lastly, participants explained that patients face barriers because they are required to obtain a prescription from a provider for injectable naltrexone prior to visiting the community pharmacy. This can not only add additional steps for the patient, but potentially for the community pharmacists.

An organizational/non-profit staff member mentioned a potential facilitator at the public policy level. They discussed the classification of OUD as a disability under the American Disabilities Act (ADA), stating that this could facilitate treatment access for formerly incarcerated individuals. However, they did not offer additional information on how this would directly apply to community pharmacist-provided naltrexone injections compared to other treatment options.

### Community Level

While discussing barriers to community pharmacist-provided naltrexone injections, many participants focused on stigma, both within the community and among providers. Stigma towards formerly incarcerated patients and treatments for substance use disorder, including OUD, can limit available services, as well as patients’ desires to seek treatment. Most of the participants noted that a major barrier is a lack of interagency collaboration between MOUD prescriber clinics, community pharmacies, and correctional institutions. Specifically, these organizations do not communicate about the healthcare status or needs of individuals transitioning back into the community, nor collaborate to facilitate treatment. Not only are releases of information an obstacle, but professionals may be unclear or make assumptions about specific responsibilities. Additionally, several participants noted that there is simply a lack of available prescribers and injectors within the community, including community pharmacists who provide injections. This not only compounds the barrier of needing to obtain a prescription, but does not provide patients with the full opportunity to utilize community pharmacists for treatment.

In terms of facilitators, only one main factor was identified at this level. A majority of participants from all stakeholder groups shared that the accessible location of pharmacies within the community could support treatment for formerly incarcerated individuals. Participants noted that this could be especially true for those who don’t have reliable transportation, as there is a good chance that there is a community pharmacy within a reasonable walking distance.

### Organizational Level

At this level, discussions focused on the community pharmacy as the organization of interest. First, several participants shared that community pharmacists may face administrative constraints to providing naltrexone injections. Specifically, they mentioned that community pharmacists can be faced with additional paperwork, and they may lack the ability to properly document individuals receiving injections. Next, a few participants shared that community pharmacies do not advertise injectable naltrexone services. This is due to deliberate choices by management or because the pharmacies lack the capacity to market their services to large audiences. The participants also added that many community pharmacists do not have the ability to provide pre-injection services, such as drug testing, or other services related to OUD treatment, including counseling or therapy. Overall, this can add additional work for community pharmacists or deter formally incarcerated individuals from using community pharmacists for injections.

When discussing facilitators, many participants representing all stakeholder groups explained that community pharmacies can offer more flexibility with appointments. Patients either don’t need to make an appointment or can easily and quickly make an appointment and often be seen the same day. Additionally, compared to other treatment options, patients have an easier time switching between community pharmacies if necessary. Also, participants added that community pharmacies can provide a nonjudgmental environment for formerly incarcerated individuals, making them comfortable enough to receive treatment at these sites. However, this category showed discordance, as a couple participants expressed concerns that they may feel judged, or the pharmacy wouldn’t provide enough privacy. One said, “The local pharmacy, these people could be, like, judgmental about a person’s addiction…So, I think the judgement could be an issue or feeling different when you walk in, you know?” (Patient/family member/caregiver). The other stated, “You’re in a public place that anyone can get into. You sit down and could have anyone you know sitting next to you…which may be a little weird now that I’m thinking about it,” (Patient/family member/caregiver). Lastly, a few participants mentioned that community pharmacies have more convenient hours than other treatment options, facilitating access for formerly incarcerated individuals. However, as with the previous facilitators, there was some disagreement among participants. In talking about pharmacy hours, a participant said, “And their hours are usually awful too. They’re not usually open. Community pharmacies at, like, [closed by] 6:30,” (Organizational staff). Another added, “So, like, with community pharmacies specifically, like, I know some of the barriers are, like, their hours. Like, they’re not usually open on Sundays, right? They have short Saturday hours. And so, for my patients that, like, again, are the off-shift workers, they can’t get their medicines from a community pharmacy,” (MOUD prescriber).

### Interpersonal Level

One main category emerged regarding barriers at the interpersonal level. MOUD prescribers and patients/family members/caregivers explained that treatment access can be hindered if formerly incarcerated individuals are released into the same home or social environment they were in prior to incarceration. They added that this often exposes these individuals to “negative” influences or temptations, causing them to fall back into old patterns of opioid use.

Compared to barriers, more facilitators were identified at this level. First, the majority of participants stated that access to naltrexone injections could be supported by patient advocates. For example, these advocates could include family members, friends, peer support specialists, or case managers. Participants added that these advocates could help keep patients accountable to their treatment schedule and goals. This facilitator was identified by participants from all stakeholder groups. Second, several participants, also representing every stakeholder group, added that if individuals have a positive, trusting, and respectful relationship with their providers, including community pharmacists, treatment is facilitated. Finally, outside of relationships, community pharmacist participants said that treatment for formerly incarcerated patients is facilitated if the pharmacy utilizes reminders via call or text.

### Individual Level

At the individual level, participants added that there is limited awareness that community pharmacists can and/or do provide naltrexone injections. Participants also noted that some patients are not only unaware that community pharmacists can provide these services, but that injectable naltrexone exists as a treatment option. Lack of awareness also prevents other non-pharmacist professionals from referring patients to community pharmacists or educating patients on this option. It also prevents reentry staff from recognizing this resource and connecting formerly incarcerated individuals to community pharmacist-provided treatment.

Additionally, the stakeholders identified several resources that create substantial barriers for formerly incarcerated individuals when not available. Nearly all participants discussed that a lack of reliable transportation – either private or public transportation – could inhibit access to naltrexone injections via community pharmacies. In addition to transportation, both a lack of insurance and lack of stable housing were identified several participants. In particular, a lack of reliable transportation was discussed by participants from all stakeholder groups.

Outside of these resources, non-patient participants described that formerly incarcerated individuals may have other responsibilities that take priority over finding and receiving treatment. Examples include finding a job, meeting with probation or parole officers, or caring for other family members, including children. These responsibilities may not only be prioritized over treatment, but create barriers for patients from a time standpoint. A few participants added that the potential side effects experienced by individuals receiving naltrexone injections, including injection site pain, may deter them from wanting to use this option.

In terms of facilitators, participants explained that treatment access is facilitated when participants have a clear plan, treatment goals, or establish their “why.” A “why” can include reasons spanning from parole requirements to being more present for family members. Finally, correctional staff and patients/family members/caregivers stated that treatment, including treatment via community pharmacies, is facilitated when individual patients are ready to make a change. This can directly relate to a patient’s “why.”

## Discussion

Overall, both barriers and facilitators were identified at every level of the Socioecological Model. In general, though, participants identified a higher number of barriers. This aligns with the idea that formerly incarcerated individuals are not utilizing community pharmacist-provided injectable naltrexone upon reentry. In terms of barriers, the most prevalent categories were at the individual level, with the public policy, community, and organizational level having an even mix. The most prevalent barrier categories included lack of interagency collaboration, inability of pharmacists to provide additional OUD services, lack of awareness, lack of insurance, and lack of reliable transportation. A focus on reducing these barriers may be an important and impactful first step in improving access to injectable naltrexone for formerly incarcerated individuals. On the other end of the spectrum, the most prevalent facilitator categories were at the organizational and interpersonal levels. These included the accessible location community pharmacies, flexibility of community pharmacy appointments, and the availability of patient advocates or social support. This not only confirms that community pharmacies are a promising resource, especially due to their accessible locations, but figuring out how to further leverage facilitators, such as patient advocates (peer support specialists, case managers, etc.), can also help improve outcomes.

Overall, there was a high level of concordance between the different stakeholder groups that participated in this study. For example, each of the categories mentioned above were identified by no less than four stakeholder groups, and most were identified by all groups. Additionally, as anticipated, formerly incarcerated individuals and family members/caregivers offered similar perspectives, supporting the decision to include these participants as one stakeholder group. There were only a few examples of discordance noted between the participants. These included discussions of community pharmacy hours and whether or not community pharmacies provide a private and non-judgmental environment for individuals to receive naltrexone injections. Participants from the individual patient/family/caregiver group in particular expressed concerns surrounding privacy and judgement, suggesting that they may have a different perception of the community pharmacy environment compared to other stakeholder groups. With that in mind, providers and support staff should not automatically assume that patients are comfortable receiving MOUD from community pharmacies, and changes to the community pharmacy environment may be necessary to ensure this comfort.

The results also showed a lot of overlap and/or connections between barriers and facilitators. First, while one participant may have identified a specific resource as a facilitator, another may have noted that the absence of that resource would create a barrier. Second, participants noted that certain factors could have an influence on each other. For example, lack of pharmacy advertising (organizational level) may directly relate to a lack of awareness of community pharmacy services (community level). Similarly, having a social support system (interpersonal level) may help an individual create a plan or identify treatment goals (individual level). Lastly, the high cost of injectable naltrexone (public policy level) may directly limit the availability of community pharmacies that provide this medication (community level).

Many of the barriers and facilitators noted by participants echo what is shown in the existing literature. This is expected, as factors impacting one MOUD option or treatment location are likely to impact access to injectable naltrexone via community pharmacies. For example, lack of insurance or lack of transportation are likely to impact treatment access, regardless of which medication or provider an individual is trying to use.^24–35^ Additionally, previous work has highlighted some of the barriers that community pharmacies face in being able to provide injectable naltrexone services, and many of these factors were identified in this study.^22^ This is also expected, as barriers to providing certain services are likely to exist regardless of the patient populations or sub-populations who may be using them. However, despite these similar findings, a significant number of categories were also specific to community pharmacist-provided treatment for formerly incarcerated individuals, and most aligned with the organizational and community level. Notably, these categories included 1) lack of interagency collaboration between primary care clinics, correctional facilities, and community pharmacies (exacerbated by patients requiring a prescription prior to injection) and 2) lack of awareness of community pharmacist-provided naltrexone services, especially among correctional staff. Community pharmacists also knew that awareness of their injectable naltrexone services was limited among other professionals and the public. They explained that this was due, in part, to the decision or inability to advertise injectable naltrexone services, which was noted as a barrier at the organizational level.

### Limitations

This study presented a few limitations that should be mentioned. First, while the researchers felt that saturation was reached and there was a high level of concordance between the different stakeholders, there were only three to four participants recruited per group. On top of that, certain participants did not have direct experience with coordinating, providing, or receiving community pharmacist-provided injectable naltrexone. These participants discussed anticipated barriers and facilitators based on their perceptions and/or experiences with community pharmacies. Also, this study did not distinguish between formerly incarcerated individuals who were released to the community from jail or prison (either with or without supervision), nor between those who were continuing or initiating injectable naltrexone upon community reentry. Overall, it is possible that saturation was not reached within each stakeholder group, or that the results may have differed with stricter inclusion and exclusion criteria as it relates to the characteristics noted above.

Additionally, several limitations relate to the transferability of the results. The stakeholders included in this study represented several counties across Wisconsin, including urban and rural areas. However, since individuals from every county could not be included, it is possible that the results are not completely representative of all stakeholders’ experiences across Wisconsin. The results may also not be generalizable to areas outside of Wisconsin. The smaller sample size also prevented the researcher from identifying urban and rural differences. Lastly, across all stakeholder groups, the participants were predominantly female, white, and did not identify as Hispanic or Latino, resulting in a homogenous sample. Despite these limitations, this study was intended to be exploratory in nature, and additional work can help ensure the transferability of these results.

### Next steps

Future research could focus on confirming these findings by including a larger sample of stakeholders or applying additional triangulation methods, such as utilizing a different framework or methodology. Additionally, as anticipated, the participants did not comprehensively discuss the barriers and facilitators that exist at the public policy level. As a result, next steps should focus on exploring the laws and regulations that impact access to community pharmacist-provided injectable naltrexone for formerly incarcerated individuals in Wisconsin. Next steps should also focus on understanding how the barriers identified in this study can be feasibly addressed through intervention or policy development, especially those that were highly prevalent and/or specific to community pharmacies. Potential interventions could focus on directly reducing barriers or helping formerly incarcerated individuals further leverage resources that support access to community pharmacist-providing injectable naltrexone. Importantly, this work can add to the current research in progress and help emphasize the importance of addressing this healthcare gap. Long-term, these findings may also be scaled-out to areas outside of Wisconsin.

## Conclusion

The barriers and facilitators identified in this study provide an opportunity to improve access to community pharmacist-provided injectable naltrexone for formerly incarcerated individuals with OUD. Overall, improving access to these services for this patient population has several social and public health implications, including decreased overdose and rearrest/reincarceration rates. Increased access can also support community health and safety and reduce existing healthcare and legal system costs. This work can also help reduce the racial and ethnic disparities that exist around this problem. Importantly, the results of this study provide a step in improving the community reentry process and ensuring that formerly incarcerated individuals with OUD are not tossed aside, but given the opportunity to receive crucial treatment and successfully reintegrate into society.

## Data Availability

All data produced in the present study are available upon reasonable request to the authors.

## Authors contributions

JC was responsible for conceptualization, data curation, formal analysis, funding acquisition, investigation, methodology, writing the original draft, and reviewing and editing. MC was responsible for conceptualization, supervision, and reviewing and editing. Both authors read and approved the final manuscript.

## Acknowledgements

The authors would like to thank James H. Ford II and Olayinka O. Shiyanbola for their contributions in conceptualizing this manuscript.

## Glossary/Abbreviations

ADA: American Disabilities Act
AODA: Alcohol and other drug abuse
DOC: Department of Corrections
MOUD: Medications for opioid use disorder
OUD: Opioid use disorder
SAMHSA: Substance Abuse and Mental Health Services Administration

